# Robust antibody levels in both diabetic and non-diabetic individuals after BNT162b2 mRNA COVID-19 vaccination

**DOI:** 10.1101/2021.07.23.21261042

**Authors:** Hamad Ali, Abdelmohsen AlTerki, Sardar Sindhu, Barrak Alahmad, Maha Hammad, Salman Al-Sabah, Mohammad Alghounaim, Mohammad H. Jamal, Ali Aldei, Mohammad J. Mairza, Maitham Husain, Sriraman Deverajan, Rasheed Ahmad, Preethi Cherian, Irina Alkhairi, Abdullah Alkandari, Jehad Abubaker, Mohamed Abu-Farha, Fahd Al-Mulla

## Abstract

The emergence of effective vaccines for COVID-19 has been welcomed by the world with great optimism. Given their increased susceptibility to COVID-19, the question arises whether individuals with type-2 diabetes mellitus (T2DM) and other metabolic conditions can respond effectively to the mRNA-based vaccine. We aimed to evaluate the levels of anti-SARS-CoV-2 IgG and neutralizing antibodies in people with T2DM and/or other metabolic risk factors (hypertension and obesity) compared to those without. This study included 262 people that took two doses of BNT162b2 (Pfizer–BioNTech) mRNA vaccine. Both T2DM and non-diabetic individuals had a robust response to vaccination as demonstrated by their high antibody titers. However, both SARS-CoV-2 IgG and neutralizing antibodies titers were lower in people with T2DM. Their levels were 154±49.1 vs. 138±59.4BAU/mL for IgG and 87.1±11.6 vs. 79.7±19.5% for neutralizing antibodies in individuals without diabetes compared to those with T2DM, respectively. In a multiple linear regression adjusted for individual characteristics, comorbidities, previous COVID-19 infection and duration since second vaccine dose, diabetics had 13.86 BAU/ml (95%CI: -27.08 to -0.64BAU/ml, p=0.041) less IgG antibodies and 4.42% (95%CI: -8.53 to -0.32%, p=0.036) less neutralizing antibodies than non-diabetics. Hypertension and obesity did not show significant changes in antibody titers. Taken together, both type-2 diabetic and non-diabetic individuals elicited strong immune responses to SARS-CoV-2 BNT162b2 mRNA vaccine; nonetheless, lower levels were seen in people with diabetes. Continuous monitoring of the antibody levels might be a good indicator to guide personalized needs for further booster shots to maintain adaptive immunity.

## 1 Introduction

COVID-19 pandemic has affected people worldwide to unprecedented proportions, even more severely affecting the people with type-2 diabetes mellitus (T2DM) and/or those with other risk factors for developing COVID-19 related complications. To date, more than 190 million people worldwide have been diagnosed with COVID-19, with an overall prevalence rate of at least 2% [1]. Although, mortality rates are generally lower (2.2%), the presence of preexisting health conditions such as T2DM, hypertension, cardiovascular disease, and metabolic syndrome may contribute to increased case fatality rates up to 10% [2; 3; 4]. A myriad of factors may contribute to increased susceptibility of T2DM patients for developing COVID-19 complications, including impaired innate/adaptive immunity, after the onset of a state of chronic, low-grade inflammation called metabolic inflammation. As a result, upon antigen exposure, the obesity-related chronic inflammation may impair macrophage activation and blunt the mechanisms of pro-inflammatory and/or innate cytokine production [5; 6]. This altered, obesogenic milieu may partly explain the presence of antiviral-resistance and vaccine escape mechanisms in obese and/or T2DM populations. Moreover, B and T cell responses are compromised in obese and more so in obese T2DM people [7]. The unfavorable hormone environment also primes for immune response dysregulation [8]. Typically, obese T2DM people may have defective innate/adaptive immunity, resulting from the enhanced production of several pro-inflammatory cytokines/chemokines like TNF-α, IFN-γ, IL-1β, IL-12, IL-18, RANTES, MCP-1 and IL-6 [9]. Indeed, all these bioactive inflammatory proteins are also the factors that have been associated with an increased susceptibility for developing COVID-19 complications [10; 11].

Driven by sheer urgency, COVID-19 vaccines have been developed at a phenomenal speed. One of the most-widely used vaccines is known as BNT162b2 (Pfizer–BioNTech) which is an mRNA-based COVID-19 vaccine that comprises of the nucleoside-modified mRNA (modRNA sequence of 4,284 nucleotides) encoding a mutated form (bases 103-3879) of the full-length spike (S) protein (peplomer) of SARS-CoV-2 stabilized in its prefusion conformation as an antigen or immunogenic molecule encapsulated in lipid nanoparticles that act as adjuvants [12]. The vaccine is administered as two shots, intramuscularly, given three weeks apart and has been shown to offer protection by triggering an immune response against infection by the SARS-CoV-2 spike protein. Since BNT162b2 vaccine delivers mRNA encoding only for SARS-CoV-2 spike protein, the expected elicited response is production of anti-S-RBD immunoglobulin G (IgG), IgM, and IgA isotypes, with neutralization potential of inhibiting the RBD binding to ACE2 cognate receptor [13].

This study aimed to evaluate the humoral immune responses in people with and without T2DM and/or other metabolic risk factors, such as hypertension and obesity. Herein, we present data showing levels of SARS-COV-2-specific IgG as well as anti-S-RBD neutralizing antibodies in a population with a high T2DM prevalence.

## 2 Methods

### 2.1 Recruitment of Participants and Study Cohort

This study was reviewed and approved by the Ethical Review Committee of Dasman Diabetes Institute “Protocol # RA HM-2021-008” as per the updated guidelines of the Declaration of Helsinki (64th WMA General Assembly, Fortaleza, Brazil, October 2013) and of the US Federal Policy for the Protection of Human Subjects. The study was also approved by the Kuwait Ministry of Health ethical committee (reference: 3799, protocol number 1729/2021). This study aimed at recruiting people that passed three weeks after taking the second dose of BNT162b2 (Pfizer–BioNTech) mRNA vaccine. People with autoimmune diseases, those taking immunosuppressants or suffering from arthritis were excluded from participating in this study as well as people with Type 1 Diabetes and pregnant women. T2DM diagnosis was based on self-reporting. Data for each participant were captured using a RedCap survey that included age, gender, existing diseases (e.g. diabetes and hypertension) as well as height, weight, and history of COVID-19 infection. Obesity was defined according to body mass index (BMI); those with BMI < 25 kg/m^2^ were considered normal weight, with BMI between 25 - 30 kg/m^2^ considered overweight and those with BMI > 30 kg/m^2^ were considered obese. Participants were then asked to visit the Dasman Diabetes Institute where they signed up the informed consent form before participating in the study.

### 2.2 Blood Sample Collection and Processing

After signing the consent form, a venous blood sample was collected in Vacutainer EDTA tubes. The blood was then centrifuged at 400 × g for 10 min at room temperature to isolate plasma. Plasma samples were then aliquoted and stored at −80°C until the assays were performed.

### 2.3 Measurement of Plasma Levels of SARS-CoV-2-specific IgG

Plasma levels of SARS-CoV-2-specific IgG antibodies were detected using enzyme-linked immunosorbent assay (ELISA) kit (SERION ELISA agile SARS-CoV-2 IgG SERION Diagnostics, Würzburg, Germany), following the manufacturer’s instructions. Briefly, plasma samples were thawed at room temperature and centrifuged for 5 min at 10,000 ×g at 4°C for sample clarification and removal of residual cells or platelets. For assay, samples were diluted 100× using sample diluent provided with the kit. Then, 100µL of controls, serum cutoffs (provided in the kit) and samples were transferred into the designated wells and incubated for 60 min at 37°C in a humid chamber. Following incubation, plates were washed 4× with 1× wash buffer and 100µL of antibody-specific enzyme conjugate was added into each well. The plate was then incubated for 30 min at 37°C in humid chamber and later washed 4× as before. Next, the chromogenic substrate (TMB solution) was added into each well and incubated for 30 min at 37°C under humidity. Finally, the reaction was stopped using the stop solution provided with the kit and the absorbance was read (Synergy H5 plate reader) to measure optical density (O.D.) at 405 nm wavelength. The test evaluation was performed following the positive and negative cutoffs as recommended by the manufacturer and IgG levels were reported as binding antibody units (BAU)/mL. In this regard, IgG levels of < 21 BAU/mL were considered negative, level of 21.0-31.5 BAU/mL were taken as border line and levels higher than 31.5 BAU/mL were considered as positive.

### 2.4 Measurement of Plasma Levels of SARS-Cov-2-specific Neutralizing Antibody

SARS-CoV-2-specific surrogate Virus Neutralization Test (sVNT) was used to detect levels of plasma neutralizing antibodies against SARS-CoV-2 S-RBD (SARS-Cov-2 sVNT kit, GenScript, USA, Inc). For sVNT assay, briefly, clarified plasma samples and kit controls were diluted 1:1 with 1× horseradish peroxidase (HRP)-conjugated recombinant SARS-CoV-2 receptor binding domain (HRP-RBD), as per the manufacturer’s instructions. Diluted samples and controls were incubated at 37°C for 30 min and later, the samples and controls, 100μL each, were transferred into designated wells in the plate and incubated for 15 min at 37°C. After incubation, wells were washed 4× with 1× wash buffer and then recommended volume of TMB solution was added into each well and again incubated for 15 min at 37°C. Finally, stop solution was added to each well and the O.D. was measured at 450 nm wavelength using Synergy H5 plate reader. The test evaluation was carried out following the recommended positive and negative cutoffs and test results were interpreted by calculating inhibition rates for samples as follows: Inhibition = (1 - O.D. value of sample/O.D. value of negative control) ×100%. According to the manufacturer’s instructions, neutralizing antibody levels higher than 20% were considered as positive.

### 2.5 Statistical analysis

We provide summary descriptive analysis using mean, median, standard deviation and interquartile range as appropriate. We fitted generalized additive linear models with plasma IgG levels and neutralizing antibodies as the dependent variables and T2DM status (yes/no), hypertension (yes/no), BMI (categorical: <25, 25-30, and >30 kg/m^2^), age, gender, previous COVID-19 infection (yes/no), comorbidity score (sum score of equal weight for heart disease, stroke, chronic obstructive pulmonary disease, asthma, obstructive sleep apnea, chronic kidney disease, bleeding disorders and other chronic diseases), and duration since receiving the second vaccine dose as the independent variables. We fitted penalized splines for the two continuous variables age and duration since second dose to account for non-linearity using restricted maximum likelihood (REML) estimation. Penalized splines are smoothing nonparametric terms that can maximize the goodness-of-fit by cross-validation and a penalty term for over- and under-fitting. We reported effect estimates with 95% confidence intervals interpreted as change in average IgG and neutralizing levels after adjustment.

In an additional *post-hoc* analysis, we also explored interaction analyses looking into the effects of being diabetic across age, gender, BMI, and hypertension categories on the circulating IgG levels. We fitted stratified adjusted models to estimate the change in IgG levels comparing diabetics to non-diabetics. We then included an interaction term between diabetes status and the other variable of interest to examine the Wald-test p-value for statistical significance. All analyses were done in R software version 3.3.1 (R Foundation for Statistical Computing), penalized splines were implemented in generalized additive models using the *mgcv* package. A p-value of less than 0.05 was considered to indicate statistical significance.

## 3 Results

### 3.1 Population characteristics

This study included a total of 262 participants;181 non-diabetics and 81 people diagnosed with T2DM. Of these 262 individuals, 193 were normotensive and 69 were hypertensive. Subdividing based on body mass index (BMI), 65 people had a BMI less than 25 kg/m^2^, 117 people had a BMI between 25-30 kg/m^2^ and 74 had a BMI higher than 30 kg/m^2^ (BMI information for 6 individuals was not available). This cohort included 126 females and 136 males **(Table.1)**.

### 3.2 SARS-CoV-2 IgG and Neutralizing Antibodies in People with or without T2DM

Analysis of the SARS-CoV-2 IgG and neutralizing antibodies based on T2DM status showed that all people elicited robust levels of these antibodies. Overall, SARS-CoV-2 IgG level was 149±52.9 BAU/mL which was more than 5 times the cut off for a positive level as recommended by the manufacturer. However, stratifying our cohort based on the T2DM status, people with T2DM had significantly lower levels of SARS-CoV-2 IgG antibodies (138±59.4 BAU/mL) compared to people without diabetes (154±49.1 BAU/mL) (**Figure 1A**). Similarly, strong SARS-CoV-2 neutralizing antibody levels were overall observed in our study cohort; however, neutralizing antibody levels were also significantly lower in people with T2DM (79.7±19.5%) compared to those without T2DM (87.1±11.6%) (**Figure 1B**). Clinical characteristics and serological findings in the studied population stratified by T2DM status are summarized in (**Table 1)**.

**Table 1.** Clinical characteristics and SARS-CoV2 serological findings in studied groups stratified by type-2 diabetes mellitus status.

|  | **Non-Diabetics**<br>**(N=181)** | **Diabetics**<br>**(N=81)** | **Overall**<br>**(N=262)** |
| --- | --- | --- | --- |
| **Age (years)** |  |  |  |
| Mean (SD) | 45.4 (14.3) | 59.4 (12.0) | 49.7 (15.1) |
| Median [Min, Max] | 43.1 [22.0, 121] | 60.3 [24.4, 81.2] | 50.3 [22.0, 121] |
| **Gender** |  |  |  |
| Female | 92 (50.8%) | 34 (42.0%) | 126 (48.1%) |
| Male | 89 (49.2%) | 47 (58.0%) | 136 (51.9%) |
| **BMI categories** |  |  |  |
| Less than 25 | 51 (28.2%) | 14 (17.3%) | 65 (24.8%) |
| Between 25 and 30 | 83 (45.9%) | 34 (42.0%) | 117 (44.7%) |
| Greater than 30 | 43 (23.8%) | 31 (38.3%) | 74 (28.2%) |
| Missing | 4 (2.2%) | 2 (2.5%) | 6 (2.3%) |
| **Hypertension** |  |  |  |
| No | 151 (83.4%) | 42 (51.9%) | 193 (73.7%) |
| Yes | 30 (16.6%) | 39 (48.1%) | 69 (26.3%) |
| **Comorbidity score** |  |  |  |
| Mean (SD) | 0.320 (0.594) | 0.469 (0.896) | 0.366 (0.703) |
| Median [Min, Max] | 0 [0, 3.00] | 0 [0, 5.00] | 0 [0, 5.00] |
| **Previous infection of COVID 19** |  |  |  |
| No | 155 (85.6%) | 69 (85.2%) | 224 (85.5%) |
| Yes | 26 (14.4%) | 12 (14.8%) | 38 (14.5%) |
| **Duration since second dose** |  |  |  |
| Mean (SD) | 84.3 (37.1) | 81.5 (37.4) | 83.4 (37.2) |
| Median [Min, Max] | 82.0 [7.00, 148] | 81.0 [13.0, 148] | 82.0 [7.00, 148] |
| **IgG (BAU/ml)** |  |  |  |
| Mean (SD) | 154 (49.1) | 138 (59.4) | 149 (52.9) |
| Median [Min, Max] | 160 [11.0, 243] | 140 [19.3, 264] | 156 [11.0, 264] |
| **IgM (BAU/ml)** |  |  |  |
| Mean (SD) | 65.6 (84.2) | 58.1 (112) | 63.3 (93.6) |
| Median [Min, Max] | 31.3 [0.600, 600] | 30.5 [0, 800] | 31.0 [0, 800] |
| **Neutralizing antibodies (%)** |  |  |  |
| Mean (SD) | 87.1 (11.6) | 79.7 (19.5) | 84.8 (14.9) |
| Median [Min, Max] | 91.5 [22.6, 95.3] | 88.0 [0, 95.8] | 90.9 [0, 95.8] |
| Missing | 3 (1.7%) | 0 (0%) | 3 (1.1%) |

**Figure 1:**
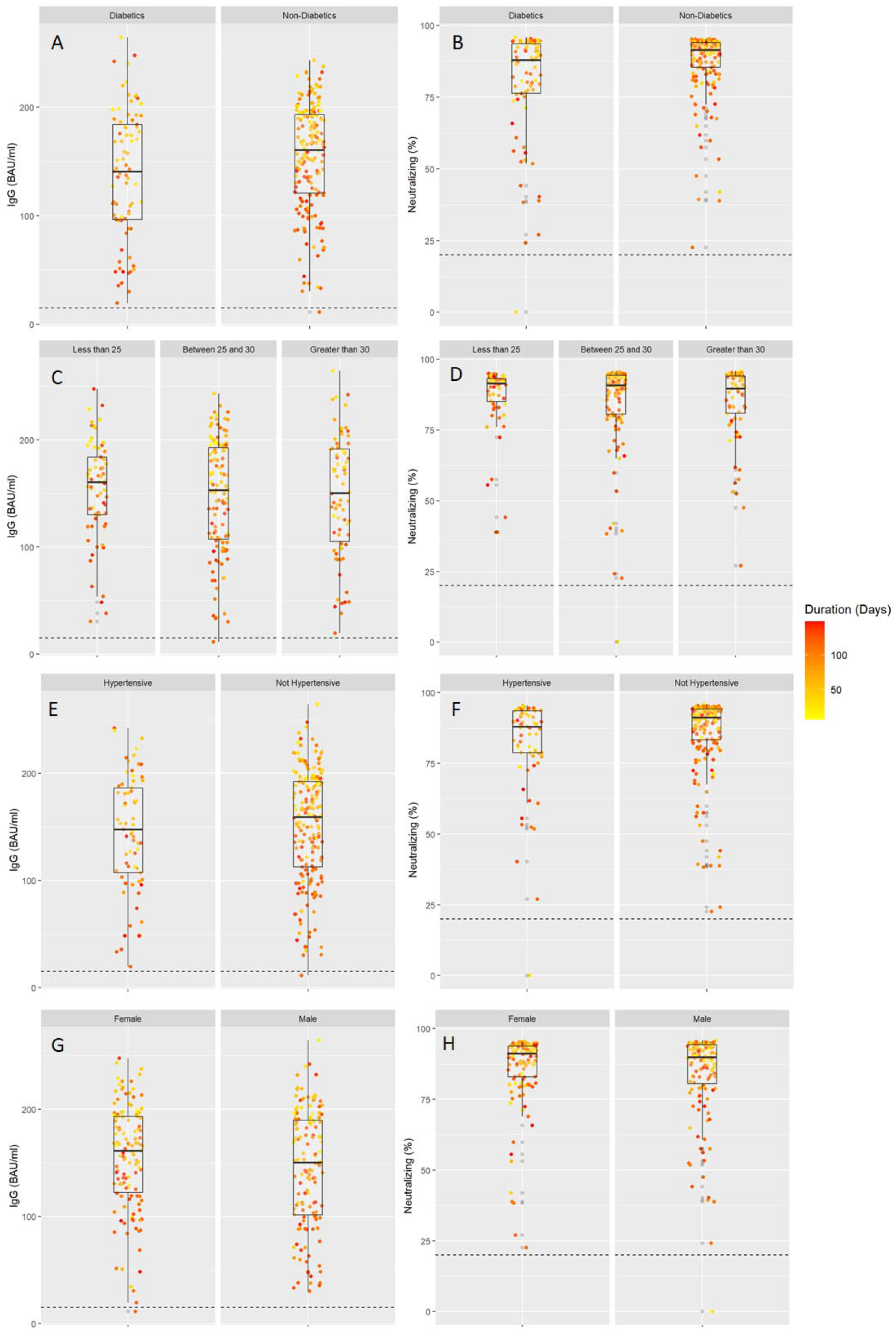
SARS-COV2 IgG and neutralizing antibodies in individuals stratified by diabetes (panels A and B), obesity levels (panels C and D), hypertension (panels E and F) and gender (panels G and H). All individuals took two doses of BNT162b2 (Pfizer–BioNTech) vaccine and this was plotted with days since vaccination shown based on the color intensity.

After adjustment to potential confounders, on average, people with TD2M had 13.86 BAU/ml (95% CI: -27.08 to -0.64 BAU/ml, p=0.041) less IgG antibodies and 4.42% (95% CI: -8.53 to -0.32%, p=0.036) less neutralizing antibodies than non-diabetics (**Table 2**).

**Table2.** Multiple linear regression analyses showing average changes of SARS-CoV2 IgG and neutralizing antibodies.

| Variable | IgG (BAU/ml)* |  |  | Neutralizing (%)* |  |  |
| --- | --- | --- | --- | --- | --- | --- |
|  | Change | 95% CI | p-value | Change | 95% CI | p-value |
| Diabetic vs. Non-Diabetic | -13.86 | [-27.08 to -0.64] | 0.041 | -4.42 | [-8.53 to -0.32] | 0.036 |
| Hypertensive vs. Not Hypertensive | 4.00 | [-9.95 to 17.95] | 0.575 | 0.75 | [-3.59 to 5.1] | 0.734 |
| Age (per 1 year increase) | -0.43 | [-0.86 to 0] | 0.049 | -0.25 | [-0.38 to -0.12] | <0.001 |
| Male vs. Female | -3.52 | [-15 to 7.96] | 0.548 | -0.31 | [-3.87 to 3.26] | 0.865 |
| BMI between 25 and 30 vs. BMI < 25 | -8.70 | [-22.58 to 5.19] | 0.221 | -1.91 | [-6.29 to 2.46] | 0.392 |
| BMI Greater than 30 vs. BMI < 25 | -5.76 | [-21.57 to 10.05] | 0.476 | -0.33 | [-5.3 to 4.63] | 0.895 |
| Previous COVID-19 Infection vs. None | 38.50 | [23.05 to 53.96] | <0.001 | 7.11 | [2.31 to 11.91] | 0.004 |
| Comorbidity score (per 1 score increase) | 4.99 | [-3.12 to 13.1] | 0.229 | 1.21 | [-1.35 to 3.77] | 0.354 |

\*All models were adjusted for the variables mentioned in addition to duration since second dose of BNT162b2 (Pfizer–BioNTech) vaccine which was modelled non-linearly with penalized splines in generalized additive models.

### 3.3 SARS-CoV-2 IgG and Neutralizing Antibodies with regard to Obesity Levels

Obesity levels also did not significantly affect the levels of SARS-CoV-2 IgG and neutralizing antibodies. The population was divided into normal weight, overweight and obese based on BMI measurements. SARS-CoV2 IgG antibody levels in people with normal weight, overweight, and obesity were 153±47.6 BAU/mL, 147.0±53.4 BAU/mL and 148.0±56.7 BAU/mL, respectively (**Figure 1C**). Similarly, SARS-CoV2 neutralizing antibody levels were also comparable in our study population and the levels in people with normal weight, overweight, and obesity were 86.3±12.8%, 84.3±16.6%, and 84.6±13.6%, respectively (**Figure 1D**). Obesity status did not show a statistically significant difference in regression analyses (**Table 2**).

### 3.4 SARS-CoV-2 IgG and Neutralizing Antibodies in People with or without Hypertension

Next, we found that the circulating levels of SARS-CoV2 IgG antibodies were comparable between people with or without hypertension. In people with hypertension, SARS-CoV2 IgG antibody levels were relatively lower (144±54.8 BAU/mL) compared to those without hypertension (151±52.2 BAU/mL) (**Figure 1E**). Similarly, neutralizing antibody levels also differed between people with hypertension and those without hypertension (81.7±17.3% vs 85.9± 13.8%) (**Figure 1F**). After adjustment to potential confounders, the differences in humoral immunity by hypertension status remained statistically non-significant (**Table 2**).

### 3.5 SARS-CoV-2 IgG and Neutralizing Antibodies by Gender, Previous COVID-19 Infection, and Age

Circulating levels of SARS-CoV2 IgG antibodies were not significantly different when compared between male and female (p=0.548) (**Table 2**). Overall, females had a higher SARS-CoV2 IgG antibody level of 154.0±50.0 BAU/mL compared to 144.0±55.2 BAU/mL in male (**Figure 1G**). Similarly, neutralizing antibody levels also differed non-significantly (p=0.865) (**Table 2**) between female and male, which were 85.9±13.5% vs 83.8± 16.0%, respectively (**Figure 1H**). Interestingly, those with the previous COVID-19 infection had higher IgG and neutralizing antibody levels (**Figure 2**) and a net average increase of 38.50 BAU/ml IgG (95% CI: 23.05 to 53.96 BAU/ml, p<0.001) and 7.11% neutralizing antibodies (95% CI: 2.31 to 11.91%, p=0.004). Regarding participant age, each one year increase in age was associated with -0.43 BAU/ml (95% CI: -0.86 to 0, p=0.049) and - 0.25% decrease in neutralizing antibodies (95% CI: -0.38 to -0.12%, p<0.001) **(Table.2)**.

**Figure 2:**
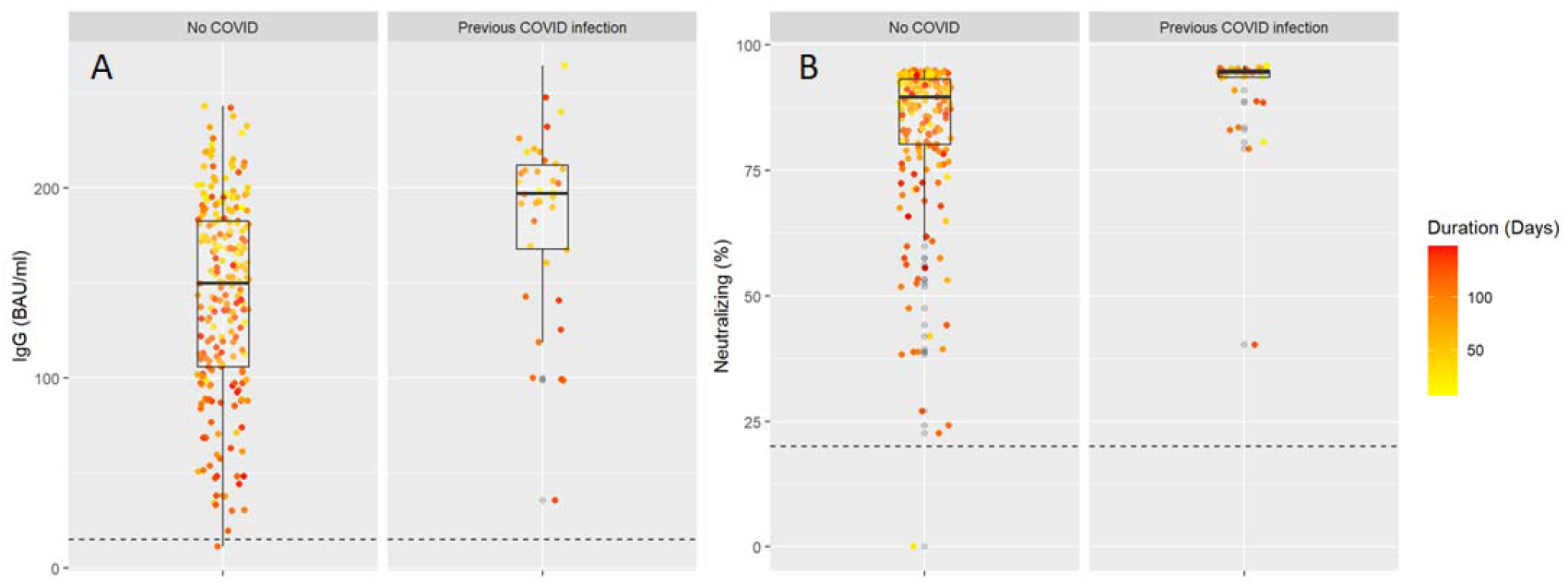
SARS-COV2 IgG (panel A) and neutralizing antibodies (panel B) in individuals with a previous COVID-19 infection. All individuals took two doses of BNT162b2 (Pfizer–BioNTech) vaccine and this was plotted with days since vaccination shown based on the color intensity.

### 3.6 Additional Analyses

Generally, IgG and neutralizing antibodies levels declined as more days passed after receiving the second vaccine dose. Antibodies-duration adjusted smooth relationships with 95% confidence intervals are presented in (**Figure 3)**. Diabetes and hypertensive status did not show considerable deviation from the linear decline over time, nor did we notice any appreciable change in slope between different subgroups. Regression estimates using IgM antibodies as the outcome also did not show any significant differences between diabetics and non-diabetics (**Supplemental Table S1**). In the interaction analyses, across all groups, being diabetic was consistently associated with lower IgG levels as compared to non-diabetics (**Supplemental Figure S1**). However, the interaction terms between diabetes status and the other variables (age, gender, previous COVID-19 infection, BMI and hypertension) were not powered to result in statistically significant p-values.

**Figure 3:**
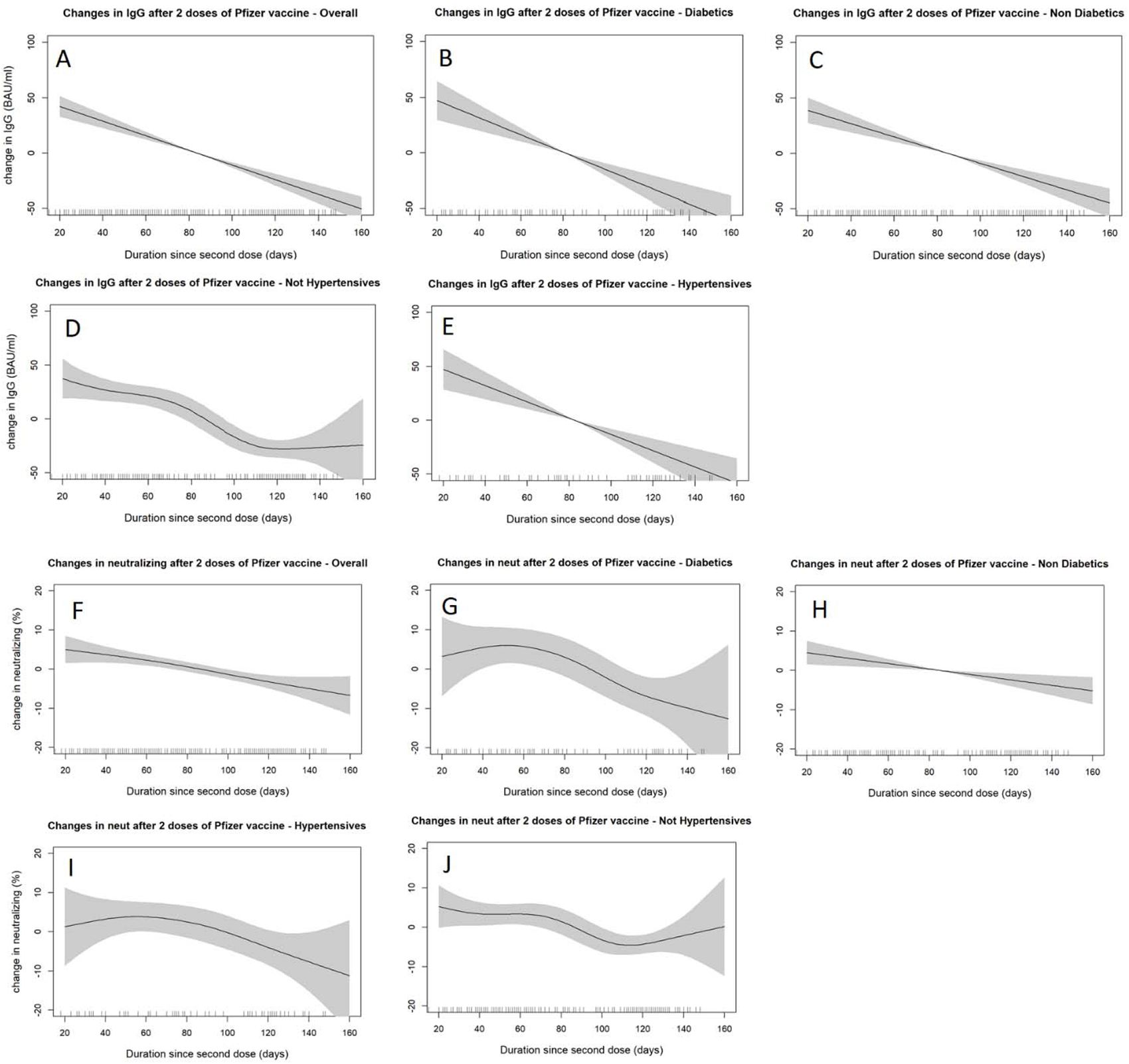
SARS-COV2 IgG (panels A-E) and neutralizing antibodies (panels F-J) decline over time since receiving the second dose of BNT162b2 (Pfizer–BioNTech) vaccine stratified by diabetes and hypertension status. The smooth relationships were derived from generalized additive models with penalized splines for duration (in days) and adjusted for age, gender, BMI, hypertension, diabetes status, comorbidity scores and previous COVID-19 infections. Solid lines represent the effect estimates of change over time while the shaded areas represent the 95% confidence intervals.

## 4 Discussion

Kuwait launched its COVID-19 mass vaccination program back in December 2020, starting from priority groups at high risk including the elderly (> 65 years), health care providers, vulnerable people with chronic underlying morbidities, members of armed forces, and others. As of now, as per the ministry of health (MOH) resources, 34% eligible people in the country have been vaccinated with at least one dose of COVID-19 vaccine and 21.26% received two doses [14]. The mass vaccination program in the country included first the BNT162b2 (Pfizer-BioNTech) mRNA vaccine and later, other COVID-19 vaccines such as adenovirus-based AstraZeneca vaccine and mRNA-based Moderna vaccine were also included. To the best of our knowledge, there has been no nationwide study looking at the humoral immune responses in the BNT162b2 recipients who have completed their COVID-19 vaccination. Therefore, in the present study, we have evaluated SARS-CoV-2-specific antibody responses after two doses of BNT162b2 (Pfizer-BioNTech) mRNA vaccine in a diverse cohort of 262 adult people, aged 49.7±15.1 years. We compared SARS-CoV-2-specific IgG and neutralizing antibody responses in these individuals with regard to T2DM status, hypertension, level of obesity, gender, and the duration after second dose.

Humoral immune response is the key component of adaptive immunity to viral infections [15]. Our data show the robust levels of SARS-CoV-2-specific IgG and neutralizing antibodies, elicited after immunization with BNT162b2 (Pfizer-BioNTech) mRNA vaccine, and titers were nearly 5 times the threshold level for seropositivity. Notably, T cell-dependent antibody response typically initiates with the production of IgM and later progresses with the expression of other immunoglobulin isotypes including IgG and IgA; all of which may mediate virus (SARS-CoV-2) neutralization [16; 17; 18]. IgM antibodies are produced early as they do not require B cells to undergo somatic hypermutation or class switch recombination, express low affinity and due to their larger size (pentamers), they are mainly found in the blood [19]. Rapid synthesis of IgM, compared to other immunoglobulin isotypes, is specifically important for infection control at the early stages via effective activation of the complement system [20]. Indeed, IgG is the major antibody isotype present in blood and extracellular fluid, while IgA is the main isotype found in secretions which makes it critical for defense against pathogens at portals of respiratory and gastro-intestinal epithelia. IgG efficiently opsonizes pathogens for removal by phagocytosis and/or by complement activation whereas IgA is a potent actor as a neutralizing antibody [21]; accordingly, IgA has been found to dominate the early neutralizing antibody response to SARS-CoV-2 infection [22; 23].

BNT162b2 vaccine is based on lipid nanoparticles formulated mRNA that encodes the full-length spike (S) protein of SARS-CoV-2 [24]. Vaccine-induced, high-affinity IgG and IgA neutralizing antibodies can inhibit the infectivity of SARS-CoV-2 by directly blocking viral attachment to host cell ACE2 receptors and preventing its entry into host cells [25; 26]. Given that BNT162b2 vaccine delivers mRNA encoding only for the SARS-CoV-2 S-glycoprotein, it would however induce the production of anti-S-RBD IgG antibodies but not anti-N IgG antibodies [27]. SARS-CoV-2 anti-N IgG antibodies are a known marker/ indicator of seroconversion after disease exposure, while the presence of SARS-CoV-2 IgG antibodies indicates seroconversion after either disease exposure or immunization [28]. Our data show robust SARS-CoV-2 IgG and neutralizing antibody levels in vaccinees after two-doses of BNT162b2 mRNA vaccine, suggesting an efficient antigen processing and presentation in all these people, which is in agreement with several other studies of BNT162b2 vaccination in diverse population cohorts [13; 29; 30].

Although, we observed high titers of anti-RBD IgG antibodies in the BNT162b2 vaccinees, these data still do not allow affirmative conclusions to be drawn as to the role of these antibodies in immunoprotection from COVID-19. The elicited anti-RBD immune response is polyclonal, nonetheless, each immunoglobulin isotype cannot be expected to play a protective role against post-vaccination SARS-CoV-2 exposure. Since there is substantial evidence for a strong positive correlation between virus neutralization assays and RBD-specific antibodies-mediated inhibition of the interaction between SARS-CoV-2 spike (S1) protein and host cell ACE2 receptor [31], we set out to measure the neutralizing antibodies that inhibited the interaction between RBD and ACE2 in a surrogate VNT and high levels of neutralizing antibodies detected in immune sera indicate that the BNT162b2 mRNA vaccine elicits protective immunity in vaccinees. In line with our findings, Pratesi et al. showed that BNT162b2 mRNA vaccine was able to induce increased levels of high-avidity, anti-RBD IgG and IgA and protective neutralizing antibodies in the vaccinated individuals [13].

Herein, we assessed whether the presence of T2DM affects the capability to mount an efficient humoral immune response against BNT162b2 mRNA vaccine. The data show that increased levels of anti-RBD IgG and neutralizing antibodies were present in both diabetics and non-diabetics. Notably, our data obtained from SARS-CoV-2 vaccinees are consistent, at least in part, with another study by Lampasona et al. reporting that humoral immune response against SARS-CoV-2 in patients with diabetes was present and superimposable, regarding timing and antibody titers, to that of non-diabetic patients [32]. However, we found that levels of RBD-specific IgG and neutralizing antibodies were significantly lower in diabetics than non-diabetics which may be explained by the fact that hyperglycemia and insulin resistance are known to induce several immune defects, such as impairment in monocyte/macrophage and neutrophil function, reduced lymphocyte proliferative responses, defective antigen presentation, and complement activation dysfunction [33]. Our findings of the lower humoral responses in vaccinees with T2DM are corroborated by Yelin et al. who found that Pfizer BNT162b2 vaccination had relatively similar effectiveness across age groups (16-80 years old), however, specific chronic comorbidities, such as T2DM, hypertension, chronic obstructive pulmonary disease (COPD), and immunosuppression were inversely associated with vaccine efficacy [34].

Next, we observed that SARS-CoV-2 IgG and neutralizing antibody levels did not differ significantly among BNT162b2 vaccinees with regard to obesity levels, age, and gender. In partial agreement with our findings, a study analyzing the antibody titers at 7 days after the second dose of BNT162b2 vaccine in a group of 248 healthcare workers reported that age and gender significantly impacted the vaccination outcome as young female participants exhibited increased humoral immune responses while BMI and hypertension did not seem to associate with difference in immune response to vaccination [35]. Pragmatically, we ought not rule out that studies of larger cohorts may unveil statistically significant differences associated with age, gender, and obesity levels.

More importantly, we found that levels of SARS-CoV-2 IgG and neutralizing antibodies were significantly higher in people who had previously had COVID-19 infection. This is comprehensible as the persons who recovered from COVID-19 infection are expected to elicit efficient humoral immunity due to the pre-existing memory B-cell responses to SARS-CoV-2. In agreement, a large cohort (n=1090) study of BNT162b2 (Pfizer-BioNTech) mRNA vaccine recipients found that the S-RBD-specific IgG and neutralizing antibody levels were higher in individuals who were previously infected with SARS-CoV-2 compared to those without prior infection. Based on these results, the authors suggested that a single dose of BNT162b2 Pfizer-BioNTech vaccine was adequate for individuals with prior SARS-CoV-2 infection, toward considering anti-S IgG antibody levels as well potential for neutralizing capability of elicited antibodies [36]. Similar findings were reported by several other studies involving diverse cohorts of various sizes [37; 38].

This study has a number of limitations. First, the population examined were self-selected mostly by word of mouth and recruitment advertisements. It is likely that the sample is over-representing co-morbid individuals who actively sought to know the antibody protection levels they have after vaccination. Second, the regression analyses adjusted for a number of *a priori* confounders, however, we cannot rule out potential residual confounding by severity of illness. For example, we could not assess whether patients with severe uncontrolled diabetes or uncontrolled hypertension show lower response to vaccine-induced humoral immunity. Third, the small sample size also limited our ability to examine the effects among different subgroups. The interaction analyses were likely underpowered to detect statistically significant differences between different strata. Fourth, the data presented represent a cross-sectional analysis of the humoral response only, therefore, further studies will be required including the longitudinal analyses of both humoral and cellular responses in the BNT162b2 vaccinees since longer-time follow-ups of both arms of the adaptive immunity can provide more precise information of the putative duration of protective immunity after vaccination. Finally, the studied population in Kuwait is likely to be homogenous and therefore the readers should be cautioned when generalizing the results to other settings and populations. We aim to further perform larger-cohort, multi-center studies analyzing both antibody persistence and presence of memory immune cell populations.

## 5 Conclusion

Taken together, our data support the presence of robust SARS-CoV-2-specific IgG and neutralizing antibody responses in people with and without T2DM, following two doses of Pfizer-BioNTech BNT162b2 mRNA vaccine against COVID-19. Notwithstanding, people with diabetes had significantly lower antibody titers than their non-diabetic counterparts; whereas, level of hypertension, obesity or gender did not show significant effect on antibody titers. Besides, continuous monitoring of SARS-CoV-2-specific IgG and neutralizing antibody profiling may be a pragmatic approach to guide personalized needs for booster shots for the maintenance of adaptive immunity in COVID-19 vaccinees.

## Data Availability

The data that support the findings of this study are available on request.

## 6 Conflict of Interest

The authors declare that the research was conducted in the absence of any commercial or financial relationships that could be construed as a potential conflict of interest.

## 7 Author Contributions

H.A., A.T. and SS Conceived and designed the analysis, researched data and wrote the manuscript. BA performed the analysis and generated the figures. M.H.1 involved in sample collection and laboratory analysis. S.S., M.G., M.J., A.A.1, A.A.2, M.M. and M.H.2 involved in subjects’ recruitment and reviewed/edited the manuscript. I.A. and P.C. performed laboratory analysis. S.D. involved in data collection and managed recruitment. R.A. edited the manuscript. J.A., M.A. and F.A. contributed to the discussion and reviewed/edited the manuscript. All authors read and approved the manuscript. M.A. is the guarantor of this work and, as such, had full access to all the data in the study and takes responsibility for the integrity of the data and the accuracy of the data analysis.

## 8 Funding

This Study was funded by Kuwait Foundation for the Advancement of Sciences (KFAS) grant (RA HM-2021-008).

## Notes

### Competing Interest Statement

The authors have declared no competing interest.

### Author Declarations

This study was reviewed and approved by the Ethical Review Committee of Dasman Diabetes Institute Protocol # RA HM-2021-008 as per the updated guidelines of the Declaration of Helsinki (64th WMA General Assembly, Fortaleza, Brazil, October 2013) and of the US Federal Policy for the Protection of Human Subjects. The study was also approved by the Kuwait Ministry of Health ethical committee (reference: 3799, protocol number 1729/2021).

