## Supplemental for "Robust antibody levels in both diabetic and non-diabetic individuals after BNT162b2 mRNA COVID-19 vaccination"

| Table S1: Change in IgM (BAU/ml) levels in adjusted multiple linear models | | | |
| --- | --- | --- | --- |
| **Variable** | **Change in IgM*** | **95% CI** | **p-value** |
| Diabetic vs. Non-Diabetic | -1.92 | [-27.07 to 23.23] | 0.881 |
| Hypertensive vs. Not Hypertensive | -13.17 | [-40.05 to 13.7] | 0.338 |
| Age (per 1 year increase) | -0.28 | [-1.12 to 0.55] | 0.508 |
| Male vs. Female | 19.28 | [-2.62 to 41.18] | 0.086 |
| BMI between 25 and 30 vs. BMI < 25 | -3.90 | [-30.82 to 23.01] | 0.776 |
| BMI Greater than 30 vs. BMI < 25 | -19.66 | [-50.1 to 10.78] | 0.207 |
| Previous COVID-19 Infection vs. None | 26.67 | [-2.7 to 56.05] | 0.076 |
| Comorbidity score (per 1 score increase) | 1.86 | [-13.67 to 17.38] | 0.815 |

***Model was adjusted for the variables mentioned in addition to duration since second dose of BNT162b2 (Pfizer–BioNTech) vaccine which was modelled non-linearly with penalized splines in generalized additive models.**

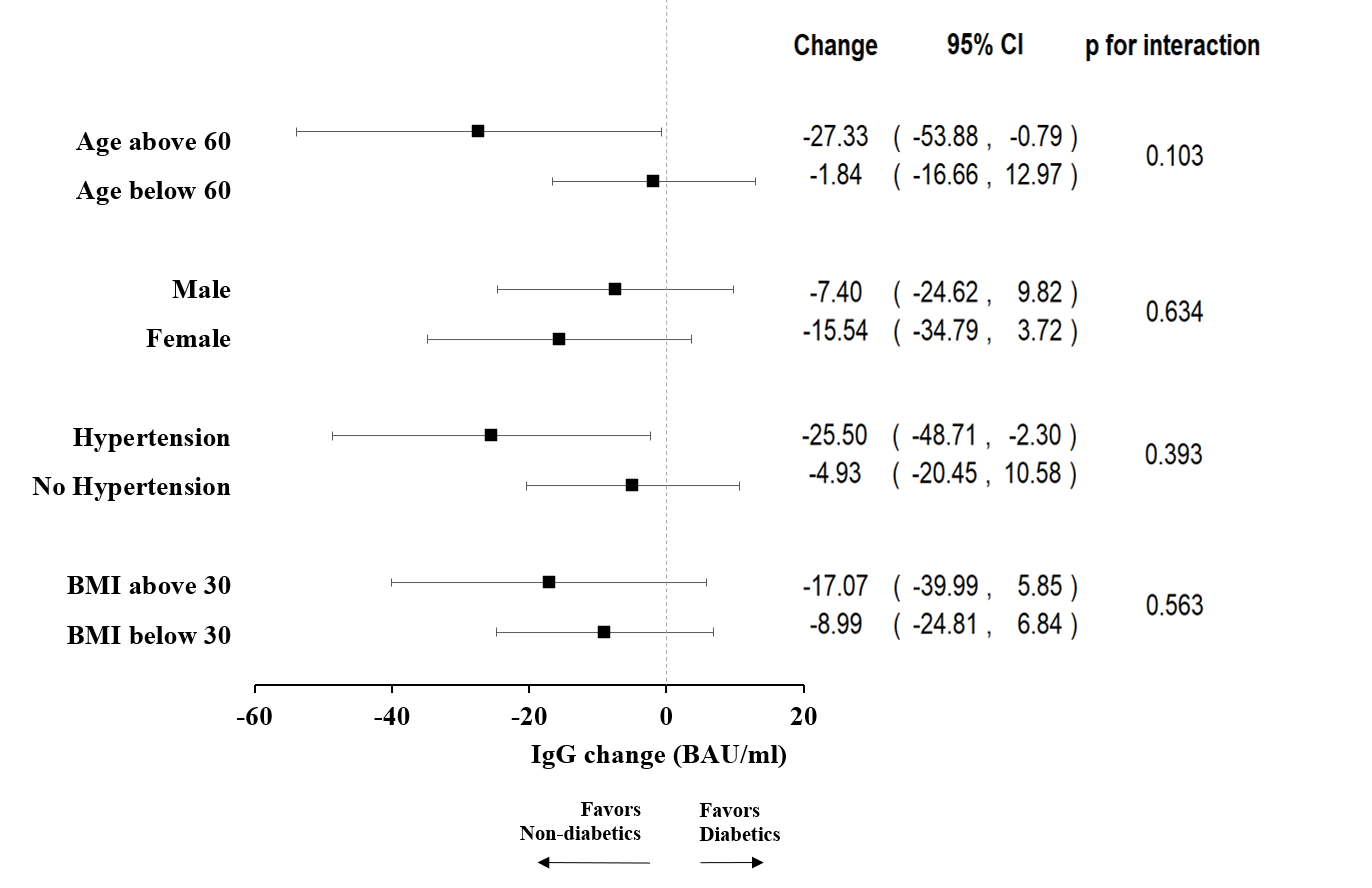

Figure S1: Forest plot showing interaction analyses of the effect of diabetes status on IgG levels (BAU/ml) stratified by age, gender, hypertension and body mass index (BMI; kg/m2). Except for the stratifying factor, all models were adjusted for age, gender, hypertension status, BMI, comorbidity score, previous COVID-19 infection and duration since the second dose of the BNT162b2 (Pfizer–BioNTech) vaccine. P-values were obtained from the Wald test after including an interaction term between diabetes status and the stratifying factor.
